# Machine Learning Assisted Web Application for Identifying Beneficial Drug Candidates for Genetic Alterations in Cancer Patients

**DOI:** 10.1101/2024.06.03.24308392

**Authors:** Hakan Şat Bozcuk

## Abstract

**Background:** Precision medicine in oncology relies heavily on molecular genetic data, primarily obtained from Next-generation sequencing (NGS) tests. However, the complexity of these data and the need to match genetic alterations with specific drug candidates pose significant challenges for clinicians. To simplify this process, a user-friendly web application has been developed. This app facilitates the matching, graphical presentation, and clustering of treatment options for specific genetic alterations, making it easier for clinicians to interpret and apply the results in patient care.

**Materials and Methods:** Utilizing the application programming interface (API) of the Drug-Gene Interaction Database (DGIdb 4.0), a web application was developed in Python to list drugs that interact with specific genetic changes. The application features a user-friendly display achieved through graphical representation and web scraping for gene-related information. Additionally, unsupervised machine learning, specifically K-means cluster analysis, was employed to categorize drug candidates based on their interaction scores with the genetic alteration in question. To enhance the interpretability of the results, the web app also provides key references and web links to the relevant drug interactions.

**Results:** The developed web application successfully filtered, listed, and displayed the gene interaction results. Utilizing an unsupervised machine learning algorithm, the app identified three optimal clusters of drug candidates based on their efficacy potentials using the Elbow Method. The cluster analysis demonstrated strong performance, evidenced by the following metrics for BRAF mutations: a Silhouette score of 0.74, a Davies-Bouldin index of 0.44, and a Calinski-Harabasz index of 475.96. Additionally, the web app effectively extracted and defined relevant gene information and identified key references for each genetic alteration within the cloud database.

**Conclusion:** The web application developed in this study provides a user-friendly platform for classifying and interpreting drug candidates based on the presence of specific genetic alterations in cancer patients. This tool is expected to enhance the accessibility and usability of genetic data, aiding clinicians in making informed treatment decisions.

## INTRODUCTION

Next-generation sequencing (NGS) is a powerful tool in genomics research, capable of sequencing many DNA fragments simultaneously. It provides detailed insights into genome structures, genetic variations, gene activity, and changes in gene behavior (1). Although next-generation sequencing (NGS) has significantly transformed the practice of oncology, interpreting NGS test results and matching these results with appropriate drug candidates remains a challenging task for clinicians. This complexity arises from the vast amount of data generated and the need for precise clinical interpretation. The transition of NGS technology from research to clinical settings has shown promise in improving cancer patient outcomes by enabling the identification of actionable mutations and personalized treatment strategies (2, 3).

NGS technology has shown improved treatment outcomes in numerous studies, particularly in advanced cancer patients, by extending progression-free survival (PFS), response rates, and overall survival (OAS) compared to control groups, although the improvements are sometimes marginal (4-9). However, some studies have reported no significant improvement in PFS with NGS-directed treatments (10). Additionally, there are barriers to accessing matched therapies. While 40% to 94% of patients undergoing NGS testing have actionable mutations, the proportion of those receiving matched therapy is significantly lower, ranging from 4% to 44% (3). Therefore, identifying accessible drug candidates and guiding oncology patients to these treatments is crucial.

It is also important to systematically search for candidate drugs for a given mutation after obtaining NGS results and extract those with a high likelihood of response. This approach can help oncologists make more informed therapeutic decisions. To address this need, this work explores the feasibility of a web application that enables systematic searches for drug candidates using cloud technologies and employs a machine learning algorithm to classify the output.

## MATERIALS AND METHODS

### General

Google Colab medium was used to develop the ML and web application software in Python coding language. A number of Python libraries and modules were utilized for the purpose of this work; “wikipedia” for accessing gene definition and function from the relevant API, “requests” as a simple and elegant HTTP library designed to handle HTTP requests, “IPython.display” for or rich media display, “sklearn.cluster” for conducting K-means clustering, “numpy” for mathematical operations, “streamlit” for web application development, and “matplotlib.pyplot” for plotting functions.

Chat GPT 4.0 was used to enhance readability, but the last version of the paper has been checked, verified, further edited and approved by the author.

### Searching gene information, genetic alteration and interacting drugs

Gene information was obtained through the Wikipedia API, to present the user a short summary of the general information about the gene structure and functions, relevant to the genetic alteration inquired. Using the Drug Gene Interaction Database (DGIdb) and by querying the API at “url = https://dgidb.org/api/graphql“, searching was conducted for the names and interaction scores of interacting drugs. Also, relevant key publications were also obtained through the PubMed API.

### Unsupervised machine learning: K-means clustering algorithm

In our study, we applied the K-means clustering algorithm, an unsupervised machine learning model, to categorize interacting drugs associated with specific genetic alterations or mutations. The primary goal of the K-means algorithm is to minimize the distance between each data point and the centroid of its assigned cluster. We utilized interaction scores, which were obtained from various cloud sources, as the key dimension for clustering. By analyzing these interaction scores, the K-means algorithm effectively grouped the drugs, providing insights into potential relationships and patterns among the drug interactions for a given genetic mutation. See Figure 1, for the working principles of the K-means clustering (11).

**Figure 1.**
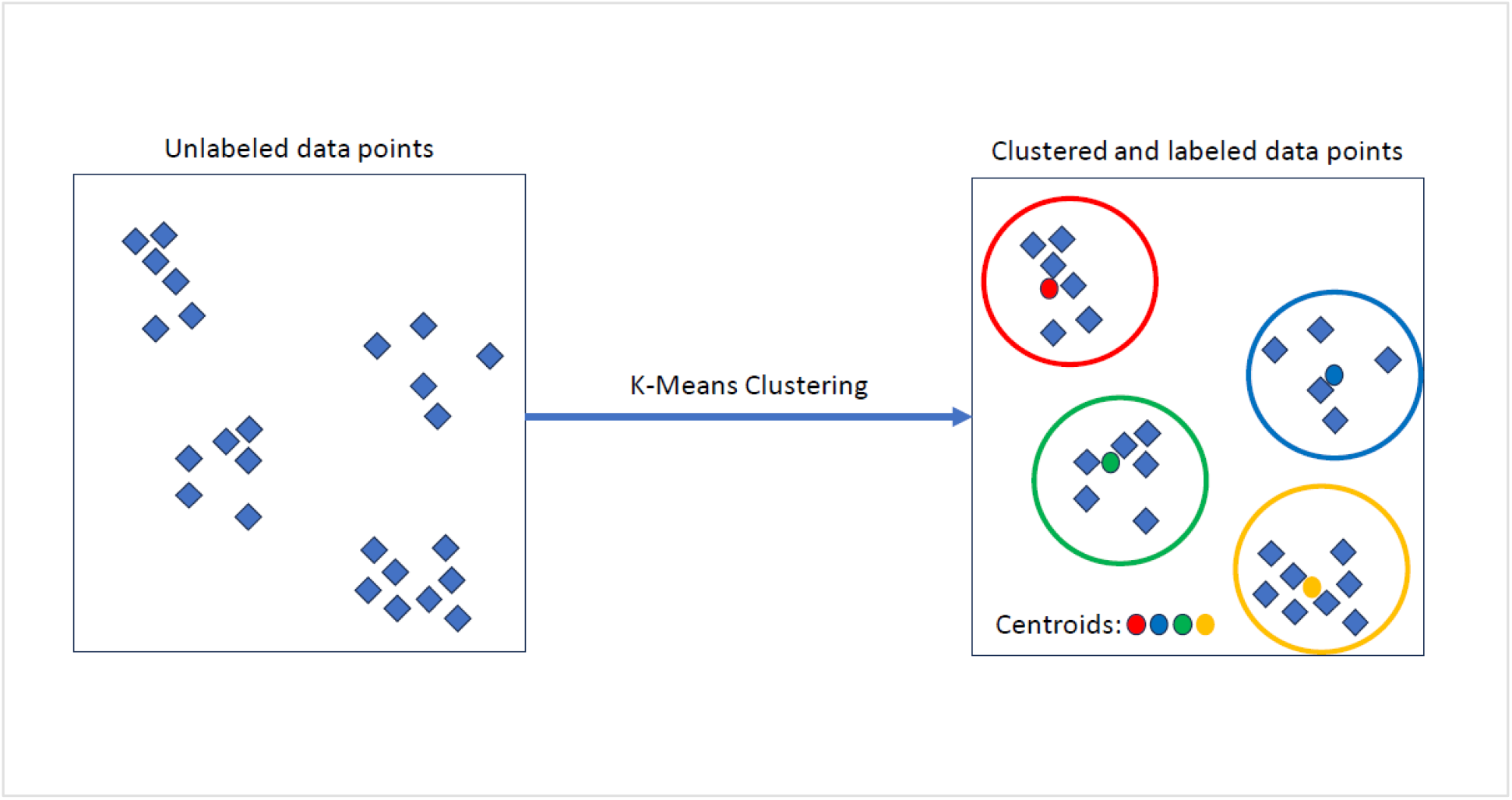
K-means clustering

In order to calculate the efficacy of the clustering algorithm efficacy, a number of measures were used; Silhouette score, Davies-Bouldin index and Calinski-Harabasz index.

The Silhouette score is a measure of how similar an object is to its own cluster compared to other clusters in clustering algorithms like K-means. It ranges from -1 to 1, where a higher value indicates better clustering. The formula for the Silhouette score for a single sample is given by:

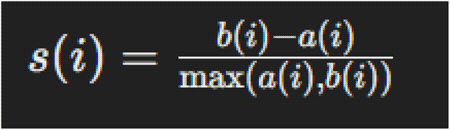

Here, a(i) is the average distance from the sample i to all other points in the same cluster, and b(i) is the minimum average distance from the sample i to all points in the nearest cluster that the sample i is not a part of (12).

The Davies-Bouldin index is a metric used to evaluate the quality of clustering algorithms, particularly in K-means clustering (13). It quantifies how well the clusters are separated and how compact they are. A lower Davies-Bouldin index indicates better clustering, with well-separated and compact clusters. The formula for Davies-Bouldin index is as follows:

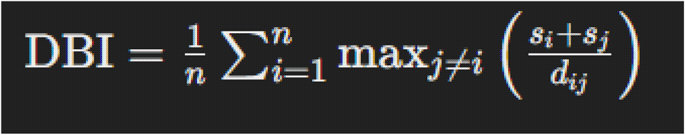

where:

- n is the number of clusters.
- s_i_ is the average distance between each point in cluster i and the centroid of cluster i (this measures the intra-cluster distance).
- D_ij_ is the distance between the centroids of cluster i and cluster j (this measures the inter-cluster distance).
- The term *max* _*j*≠*I*_ (s_i_ + s_j_) / (d_ij_) calculates the worst-case ratio of intra-cluster distance to intercluster distance for each cluster.

The Calinski-Harabasz index, also known as the Variance Ratio Criterion, is a metric used to evaluate the quality of clustering in a dataset (14). It measures the ratio of the sum of between-cluster dispersion and of within-cluster dispersion for all clusters. A higher Calinski-Harabasz index indicates a better-defined clustering. Calinski-Harabasz index is calculated according to the following formula:

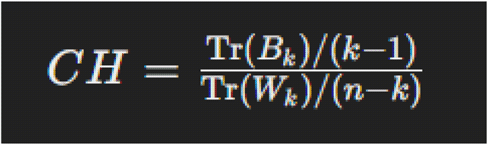

where:

- Tr(B_k_) is the trace of the between-cluster dispersion matrix.
- Tr(W_k_) is the trace of the within-cluster dispersion matrix.
- k is the number of clusters.
- n is the total number of data points.

The between-cluster dispersion matrix B_k_ measures how far the cluster centers are from the overall center of the dataset, while the within-cluster dispersion matrix W_k_ measures the compactness of the clusters.

The optimal number of clusters was tested on a large number of genes with the use of Elbow method. Appropriate plots were drawn to make use of the Elbow method for different genes.

### Plotting the results

Two figures were generated for each genetic alteration query. Firstly, the top 10 drugs with the highest interaction scores relevant to a genetic alteration were plotted. Secondly, 3 clusters of interacting drug candidates with reference to their interaction scores were plotted. These clusters showed drugs with different activities; i.e. high, medium and low interaction scores, for a given genetic alteration.

### Development of web app

The web application is developed again in Python language, using Spyder medium, saved in GitHub platform, developed by the Streamlit library, and deployed on Render platform. The resultant web app enabled search for genetic alterations, outputting interacting drugs, their ML clusters, general gene information, key references, with various plots.

## RESULTS

### General appearance and functionality of the web app

DGIdb contains over 10,000 genes and 15,000 drugs involved in over 50,000 drug-gene interactions or belonging to one of 43 potentially druggable gene categories (15,16).

Web app can be assessed on any digital tool with a web browser. Gene interactions viewer with its side panel where brief information about the web app is presented, and the gene information text output can be seen in Figure 2, from 2a to 2c.

**Figure 2.**
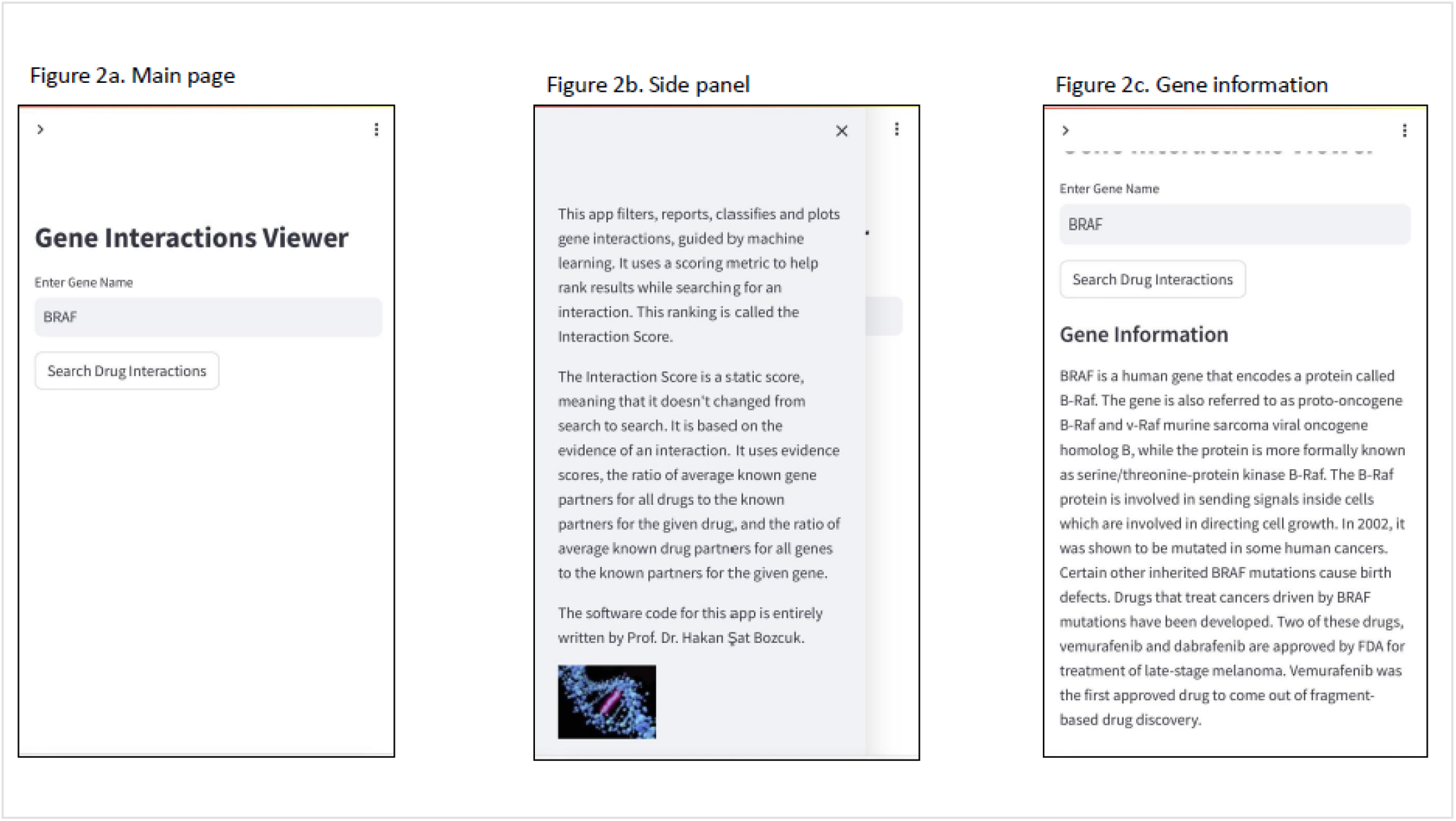
Web app: main page and general information

Likewise, a plot that is produced after the search button is pressed and which shows the first 10 drug interactions for a given genetic alteration can be viewed in Figure 3a. The second plot presents the drug clustering results, as seen in Figure 3b. Of note, drug interactions and the clusters they belong to are listed below the clustering plot; an example search for BRAF yields the list in Figure 3c. Lastly, key references about the candidate drugs are detailed at the last part of the output, with links to the original PubMed publications, as demonstrated in Figure 4.

**Figure 3.**
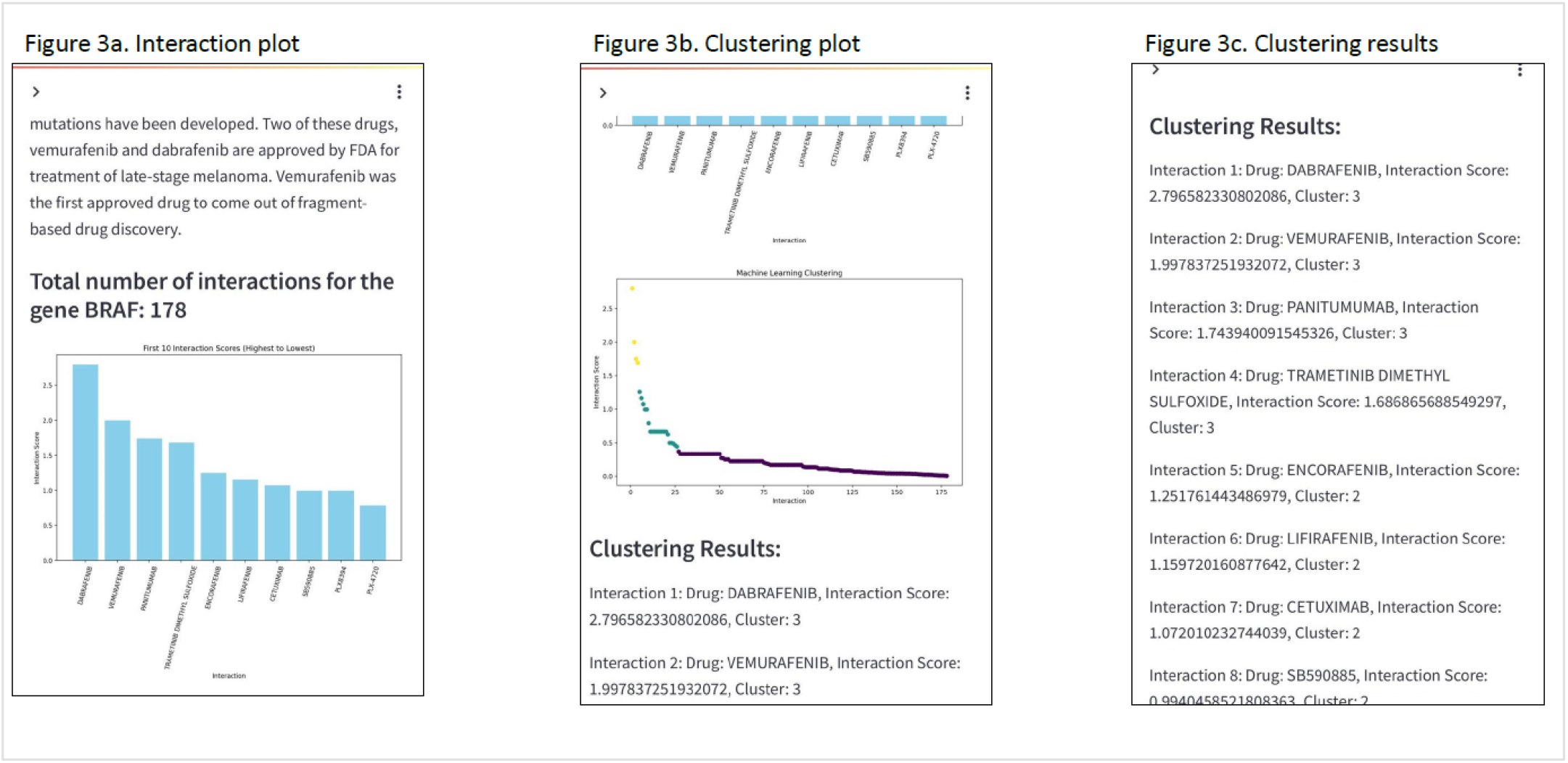
Web app: main plots and clustering results

**Figure 4.**
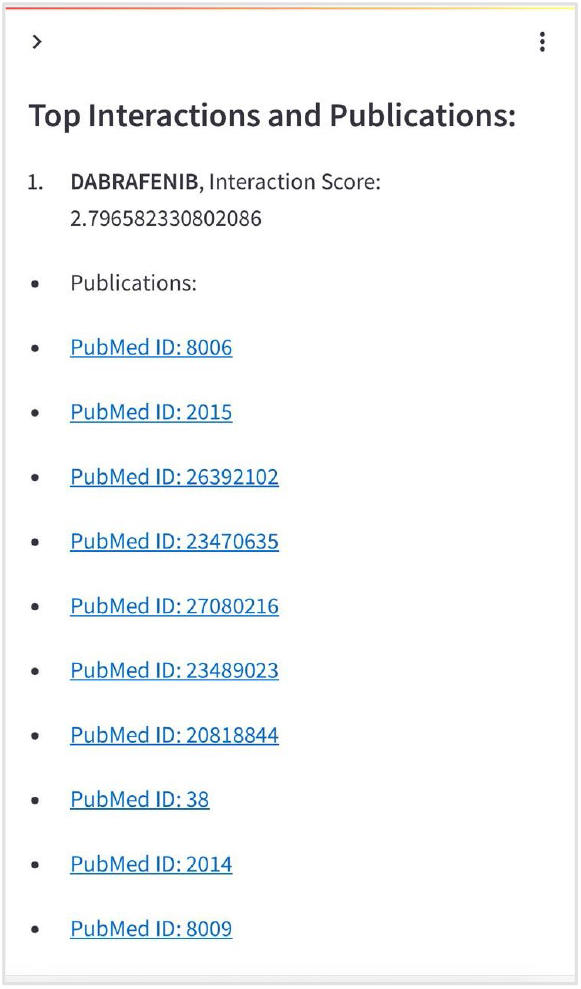
Web app: references

### Machine Learning and Cluster Analysis results

3 genomic targets were randomly chosen among around the 10000 genes in DGIdb to represent the distribution of clustering efficacy measures for different genomic alterations, and these targets were BRAF, EGFR and PTEN. Elbow method and its associated plot helped identify the optimal number of clusters of gene interactions; i.e. drug candidates. Figure 5 shows the optimal number of clusters for EGFR interactions is 3.

**Figure 5.**
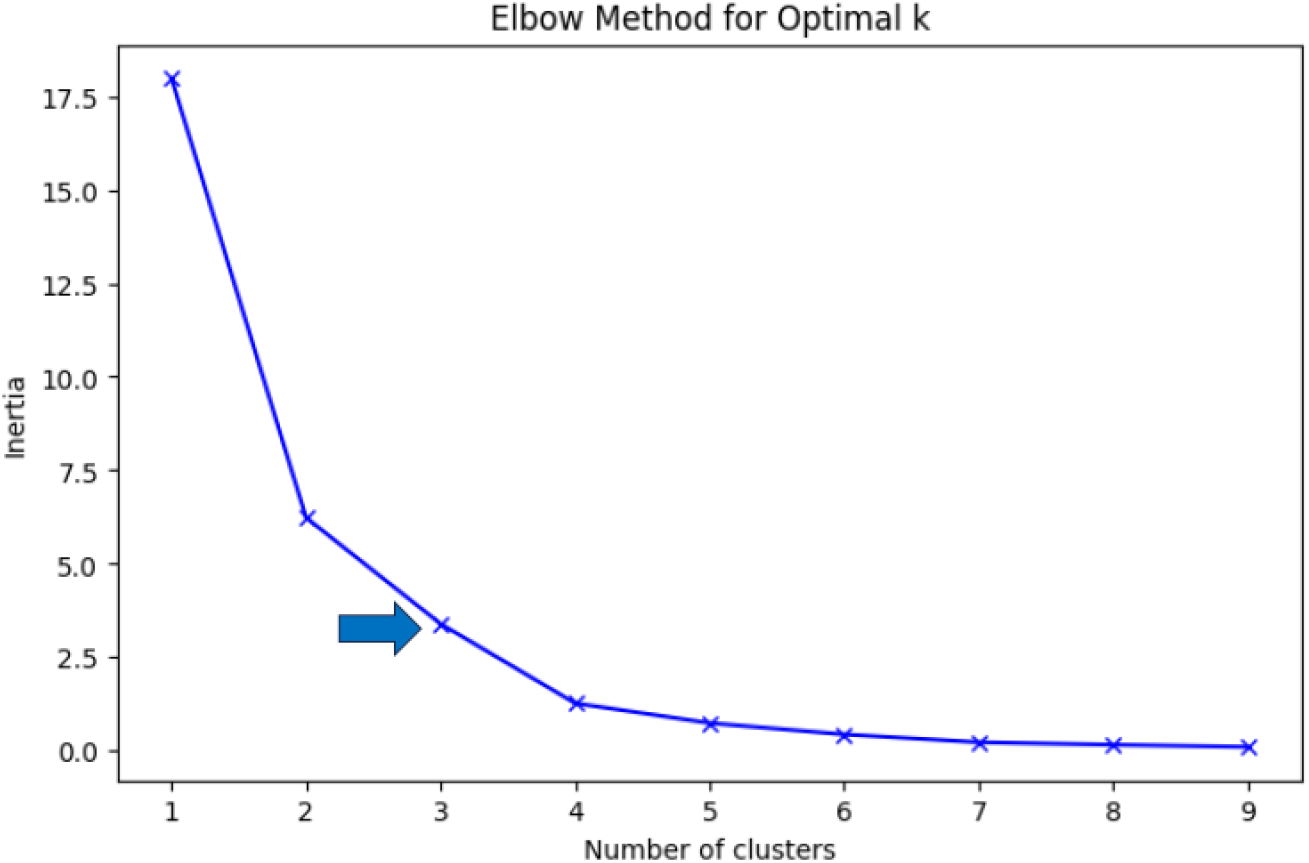
Elbow method of calculating optimal cluster numbers

K-Means Cluster analysis of EGFR yielded the following 3 clusters among the 185 interactions, as seen in Figure 6.

**Figure 6.**
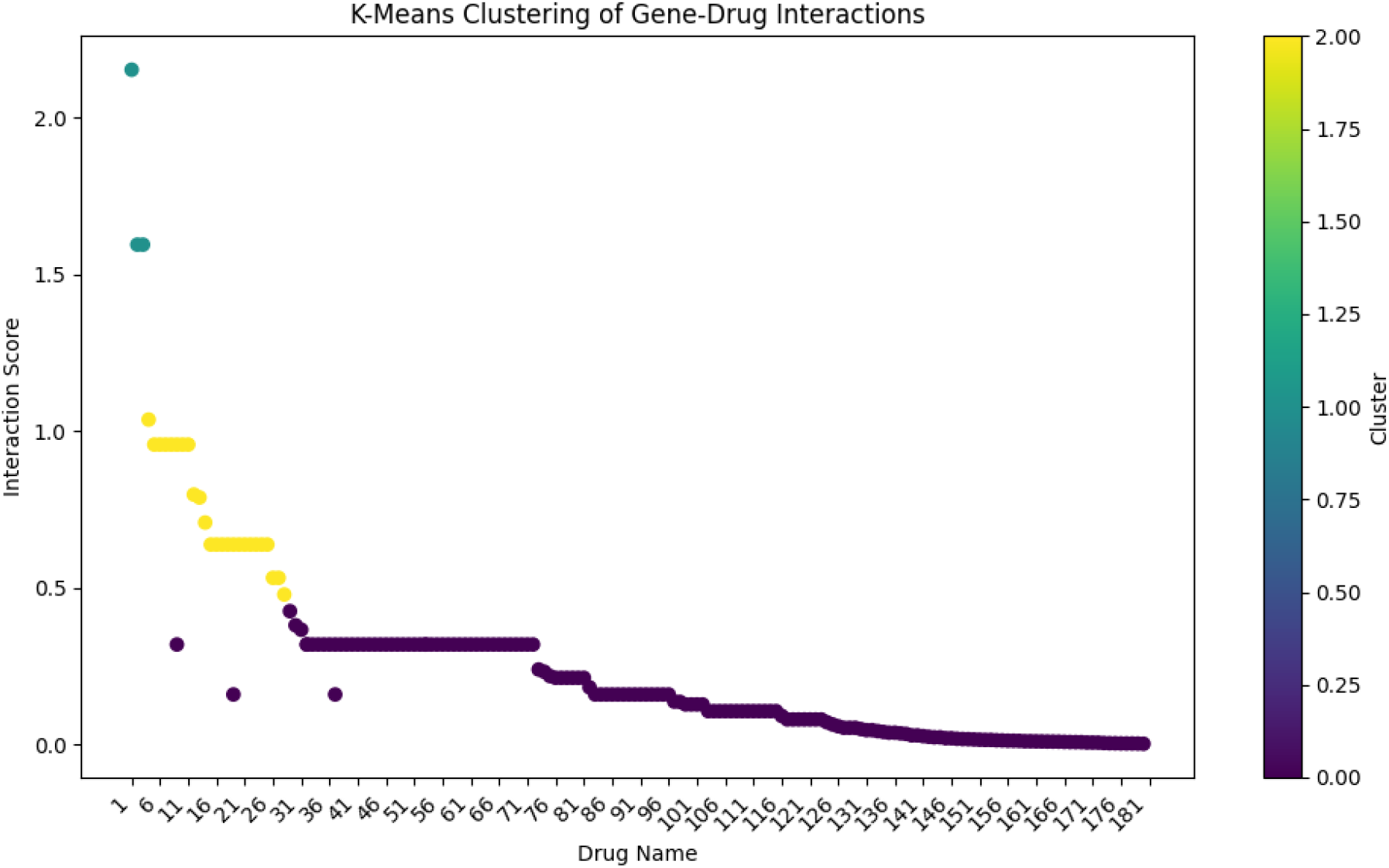

For clustering efficacy metrics for the 3 genes, refer to Table 1 for details.

**Table 1.** Clustering efficacy metrics for 3 genes.

| Gene | Total no of genetic interactions | Silhouette score | Davies-Bouldin index | Calinski-Harabasz index | Optimal number of clusters* |
| --- | --- | --- | --- | --- | --- |
| BRAF | 178 | 0.74 | 0.44 | 475.96 | 3 |
| EGFR | 185 | 0.73 | 0.42 | 390.59 | 3 |
| PTEN | 145 | 0.74 | 0.34 | 480.32 | 3 |

\*; Calculated according to the Elbow method.

## DISCUSSION

The findings of this study demonstrate that a user-friendly web application leveraging machine learning to classify treatment options into clinically meaningful clusters is feasible for oncology. Furthermore, such an application is expected to enhance understanding for both clinicians and patients, thereby improving treatment decisions following Next-Generation Sequencing (NGS) testing. A similar application has been developed for psychiatry, where there are fewer genetic alterations and drug options, and has proven to be suitable for clinical practice. (17)

The development and deployment of a machine learning-assisted web application for identifying beneficial drug candidates for genetic alterations in cancer patients represents a potential for significant advancement in precision oncology. Precision medicine relies heavily on molecular genetic data obtained from Next-generation sequencing (NGS) tests, which can identify actionable mutations to guide treatment strategies. However, the interpretation of these results and matching them with suitable drug candidates remains a challenge due to the complexity and volume of data generated (18). This study addresses this challenge by creating a user-friendly web application that integrates data retrieval, graphical representation, and machine learning clustering to aid clinicians in making informed treatment decisions.

The integration of the Drug-Gene Interaction Database (DGIdb) with machine learning algorithms provides a robust framework for identifying and categorizing drug interactions based on their efficacy. The application of the K-means clustering algorithm enabled the grouping of interacting drugs into categories, which were validated using various clustering efficacy metrics such as the Silhouette score, Davies-Bouldin index, and Calinski-Harabasz index. These metrics indicated strong clustering performance, with scores reflecting well-defined and compact clusters (12, 13). For example, the Silhouette score of 0.74 for BRAF mutations suggests a high degree of separation between clusters, while the Davies-Bouldin index of 0.44 indicates minimal overlap between clusters. The strong performance of the clustering algorithm is anticipated to provide a clinically valuable method for classifying drug candidates into categories of good, medium, and poor efficacy for a specific genetic alteration.

The graphical representation of drug interactions and the clustering results enhances the interpretability of the data for clinicians. By visualizing the top interacting drugs and their clusters, clinicians can quickly identify potential treatment options and their relative efficacy. This approach is supported by literature, which emphasizes the importance of visual aids in clinical decision-making and data interpretation (19, 20). Moreover, the inclusion of key references and links to PubMed publications provides an additional layer of validation and credibility to the results, facilitating further exploration of the identified drug candidates.

Furthermore, the use of cloud technologies and APIs for data retrieval and processing ensures that the web application remains scalable and up-to-date. The reliance on cloud sources for interaction scores and gene information allows for real-time updates and access to a vast repository of genomic data (21). This capability is crucial for keeping pace with the rapid advancements in genomics and the ever-expanding list of known genetic alterations and their associated drug interactions.

In conclusion, the developed web application not only simplifies the process of matching NGS results with appropriate drug candidates but also provides a reliable and efficient tool for precision oncology. The integration of machine learning clustering enhances the ability to categorize drug interactions, aiding in the identification of the most promising treatment options. Future directions for this work could include the incorporation of additional data sources, such as clinical trial results, to further refine and validate the drug interaction predictions. This would enhance the utility of the application and support its relevance in the evolving landscape of precision medicine.

## Data Availability

All data produced are available online at “https://www.dgidb.org/”.

https://www.dgidb.org/

## Acknowledgement

All the code associated is the sole work of the author, and also the web application has been developed and coded exclusively by the author.

